# Comparative Performance of an AI Tool and First-Year Residents for Retinal Disease and Glaucoma Assessments: A Study in a Mexican Tertiary Care Setting

**DOI:** 10.1101/2024.08.26.24311677

**Authors:** Dalia Camacho-García-Formentí, Gabriela Baylón-Vázquez, Karen Arriozola-Rodríguez, Enrique Avalos-Ramirez, Curt Hartleben-Matkin, Hugo Valdez-Flores, Damaris Hodelin-Fuentes, Alejandro Noriega

## Abstract

**Background:** Artificial intelligence (AI) shows promise in ophthalmology, but its potential on tertiary care settings in Latin America remains understudied. We evaluated a Mexican AI-powered screening tool, against first-year ophthalmology residents in a tertiary care setting in Mexico City.

**Methods:** We analysed 435 adult patients undergoing their first ophthalmic evaluation. AI and residents’ assessments were compared against expert annotations for retinal disease, cup-to-disk ratio (CDR) measurements, and glaucoma suspect classification. We also evaluated a synergistic approach combining AI and resident assessments.

**Results:** For glaucoma suspect classification, AI outperformed residents in accuracy (88.6% vs 82.9%, *p* = 0.016), sensitivity (63.0% vs 50.0%, *p* = 0.116), and specificity (94.5% vs 90.5%, *p* = 0.062). The synergistic approach deemed a higher sensitivity (80.4%) than ophthalmic residents alone or AI alone (*p <* 0.001). AI’s CDR estimates showed lower mean absolute error (0.056 vs 0.105, *p <* 0.001) and higher correlation with expert measurements (*r* = 0.728 vs *r* = 0.538). In retinal disease assessment, AI demonstrated higher sensitivity (90.1% vs 63.0% for medium/high-risk, *p <* 0.001) and specificity (95.8% vs 90.4%, *p <* 0.001). Furthermore, differences between AI and residents were statistically significant across all metrics. The synergistic approach achieved the highest sensitivity for retinal disease (92.6% for medium/high-risk, 100% for high-risk).

**Conclusion:** AI outperforms first-year residents in key ophthalmic assessments. The synergistic use of AI and resident assessments shows potential for optimizing diagnostic accuracy, highlighting the value of AI as a supportive tool in ophthalmic practice, especially for early-career clinicians.

## INTRODUCTION

The need for ophthalmic screenings in Mexico has increased significantly due to high prevalence of risk factors associated with ophthalmic diseases. Diabetes, which affects approximately 15 million Mexicans over 20 years old [1–3], is a risk factor for several conditions. These include glaucoma, diabetic retinopathy (DR), diabetic macular edema (ME), and cataracts [4–6]. Furthermore, the elderly population also faces higher risks of glaucoma, age-related macular degeneration (AMD) and cataracts [4, 6, 7].

Glaucoma presents additional risk factors, including family history of glaucoma and increasing age [4], affecting primarily individuals over 40 years old [8]. It is estimated that over 1.5 million Mexicans have glaucoma and 50% remain undiagnosed [9]. Even though, glaucoma is the second leading cause of irreversible blindness in Mexico and worldwide [9].

While periodic ophthalmic evaluations are recommended for these at-risk populations, there is a scarcity of ophthalmologists. For example, by July 2024 there were only 4,213 ophthalmologists registered in Mexico, 31.5% of them in Mexico City [10]. To address the growing need of ophthalmic screenings worldwide, researchers have developed AI models for retinal disease screenings using fundus images, [11–16] as well as models that analyse the optic disk for glaucoma suspect classification [17–19]. Some AI systems and models have been validated in Latin America [12, 13, 16].

For example Arenas-Cavalli *et al* have validated DART, a DR screening tool, on the Chilean health system [16] and González-Briceño *et al* evaluated their models on primary care data from the Mexican Institute of Social Security [12].

These validations were focused mainly on DR screening on primary care settings. To our knowledge, AI systems that identify a broader range of retinal diseases, as well as glaucoma suspects have not been evaluated in Latin America. Furthermore, their potential in tertiary care settings remains unexplored, even though experienced ophthalmologists show considerable variability in CDR estimation and retinal disease assessments. [17, 20–22]. Furthermore, previous studies have demonstrated that AI can enhance ophthalmologists’ sensitivity for DR classification [23], and AI-based CDR measurements have surpassed the average expert [17].

For early-career clinicians AI can improve the learning process for retinal disease classification [24, 25]. This potential for AI to support early-career clinicians is also valuable in clinical practice, considering that first-year residents are often in charge of first-time consultations at ophthalmology hospitals in Mexico.

Considering this, we evaluated the performance of an AI-powered ophthalmic screening tool during first-time consultations at a Mexican ophthalmology hospital. Our evaluation focused on CDR estimation, glaucoma suspect and retinal disease classification. We compared the AI’s performance to that of first-year ophthalmology residents. Additionally, we explored a synergistic approach between AI and residents, highlighting how AI could support early-career clinicians in assigning diagnoses and referring patients to subspecialty consultations.

Compared to previous research, our study evaluates the specific use case of AI in an ophthalmology hospital in Latin America, and compares it against first-year ophthalmology residents, which is the standard benchmark in ophthalmology hospitals in Mexico for first-time consultations. By evaluating AI in this setting, we assess the possible improvement that could be achieved on patient care at ophthalmology hospitals.

## MATERIALS AND METHODS

We performed screenings with an AI-based ophthalmic screening tool on patients over 18 years old, that underwent their first ophthalmic evaluation by a first-year ophthalmology resident. These studies were carried out from February 12 to March 14, 2024, at Conde de Valenciana Centro, an ophthalmic institute in Mexico City.

This study was conducted according to the Declaration of Helsinki guidelines and was approved by Conde de Valenciana’s Ethics in Research Committee (CEI-2023/12/01), Biosecurity Committee (CB-0053-2023), and Research Committee (CI-053-2023).

For each patient, we collected the hospital medical record ID, personal information, information related to risk factors, and ophthalmic symptoms. The complete list of variables is presented on Appendix A.

Retinal fundus images were also required for screening, these were taken with a Horus 45° autofocus portable fundus non-mydriatic camera from Jedmed.

### Data

A total of 464 patients were screened, nine screenings were excluded from the analysis due to registration errors in the AI platform. Six studies were excluded because of missing images resulting from camera problems. Additionally, 14 patients were excluded, due to empty medical records. Thus, 435 screenings were considered for analysis. The average age was 59.1 (15.7 SD) years, 34.0% of patients were men and 66.0% were women, 32.2% reported diabetes and 39.3% hypertension.

### Medical records

Medical records of the patients involved in this study were identified by the medical record ID. We extracted the following information: CDR by eye, initial diagnosis defined by the first-year residents and whether it was associated with glaucoma, retina, cataract or another subspecialty.

### AI screening tool

The screenings for this project were conducted using retinIA (v 3.3.1), an AI-based ophthalmic screening tool developed by PROSPERiA [26].

Compared to other screening tools, such as EyeArt, DART, and Retinalyze, which focus primarily on DR, AMD, or glaucoma, retinIA provides a more comprehensive analysis [11,16,27], identifying a broader range of retinal diseases including DR, ME, AMD, and PM, as well as presence of other retinal disease and risk of visual loss. It also provides CDR estimation for glaucoma suspect classification, identifies possible media opacities, other ophthalmic conditions, and provides explainability features. Appendix B provides further details on the underlying AI models.

### Synergistic Approach

The synergistic approach for both glaucoma suspects and retinal disease assessments considers the outcome to be positive if either the residents or the AI tool determined a positive outcome.

### Ground-truth annotation

A total of 1,013 fundus images were obtained during data collection, 918 belonged to the 435 patients considered for analysis.

To determine the ground truth value for CDR analysis, three ophthalmologists annotated 861 images, where the optic disk was present, using LinkedAI annotation platform [28]. The ground truth CDR was calculated as the average CDR between the three experts.

We determined ground truth for glaucoma suspect as follows. If CDR was 0.6 or more, or if the CDR of one eye exceeded the other by 0.2 or more, the patient was then considered as a glaucoma suspect. These criteria were based on Harizman et al’s definition on absence of glaucomatous optic neuropathy, considering measurements associated to CDR [29].

To determine ground truth for presence of retinal disease, all 918 images were annotated by a retina expert and an ophthalmic expert. For images in which there was disagreement between experts, ground truth was obtained through negotiation. Annotation of retinal findings was performed on Televal platform [30]. A complete list of all annotated findings can be found in Appendix C and the rules to determine prediagnosis and risk of visual loss are presented in Appendix D.

## RESULTS

For both glaucoma suspects and presence of retinal disease, we calculated accuracy, specificity, sensitivity, positive predictive value (PPV) and F1-score. To account for statistically significant differences between AI and residents, we calculated p-values using bootstrap estimation method. For glaucoma suspects we also calculated the receiver operating characteristic curve (ROC) considering the maximum estimated CDR by patient as the model. For CDR we evaluated the absolute and relative errors and calculated the Pearson correlation coefficient (*r*). An additional comparison between cataract diagnosis and media opacities is shown in Appendix E.

### Glaucoma suspect and cup disk ratio

The AI software analysed CDR for 61.6% of patients, for the remaining 38.4% the software considered patients’ images had insufficient quality for CDR analysis. Meanwhile, ground truth CDR was obtained for 78.6% of patients. On the other hand, residents, who evaluate patients at an in-person consultation, registered CDR for 95.4% of patients. The analysis to assess performance was done on 245 patients (56.3%), corresponding to those who had a CDR evaluation by the residents, the AI tool, and all three ophthalmologists. Metrics for glaucoma suspect classification are shown in Table 1. For AI, patients were classified as glaucoma suspects if the estimated CDR was 0.55 or higher. This threshold was set by maximizing the F1-score.

**Table 1:** Performance metrics for glaucoma suspect classification. The values considered for AI correspond to a high specificity point, where the best F1-sccore is met.

|  | **Accuracy** | **Sensitivity** | **Specificity** | **PPV** | **F1 score** |
| --- | --- | --- | --- | --- | --- |
| **Resident**<br>**diagnosis** | 82.9%<br>(78.0%,<br>87.3%) | 50.0%<br>(35.3%,<br>64.8%) | 90.5%<br>(86.2%,<br>94.1%) | 54.8%<br>(39.1%,<br>69.7%) | 52.3%<br>(38.0%,<br>64.5%) |
| **AI** | **88.6%**<br>(84.5%,<br>92.2%) | 63.0%<br>(50.0%,<br>77.3%) | **94.5%**<br>(91.1%,<br>97.6%) | **72.5%**<br>(57.1%,<br>86.1%) | **67.4%**<br>(55.2%,<br>77.8%) |
| **Resident**<br>**or AI** | 85.3%<br>(80.4%,<br>89.4%) | **80.4%**<br>(68.3%,<br>91.5%) | 86.4%<br>(81.2%,<br>90.7%) | 57.8%<br>(44.9%,<br>69.6%) | 67.3%<br>(55.4%,<br>76.0%) |

When comparing AI’s performance to that of the ophthalmology residents, AI consistently outperformed residents across all metrics. The AI tool achieved an accuracy of 88.6% compared to 82.9%, a sensitivity of 63.0% compared to 50.0%, a specificity of 94.5% compared to 90.5%. Statistically significant differences were found for accuracy (p = 0.016), PPV (p = 0.02), and F1-score (p = 0.026). Differences in sensitivity (p = 0.116) and specificity (p = 0.062) were not statistically significant. The synergistic approach yielded the highest sensitivity (80.4%) and differed significantly from both AI (p < 0.001) and resident’s performance (p < 0.001).

In Figure 1 we show the ROC curves for AI and residents of ophthalmology considering the maximum CDR by patient as the model for both AI and the residents.

**Figure 1:**
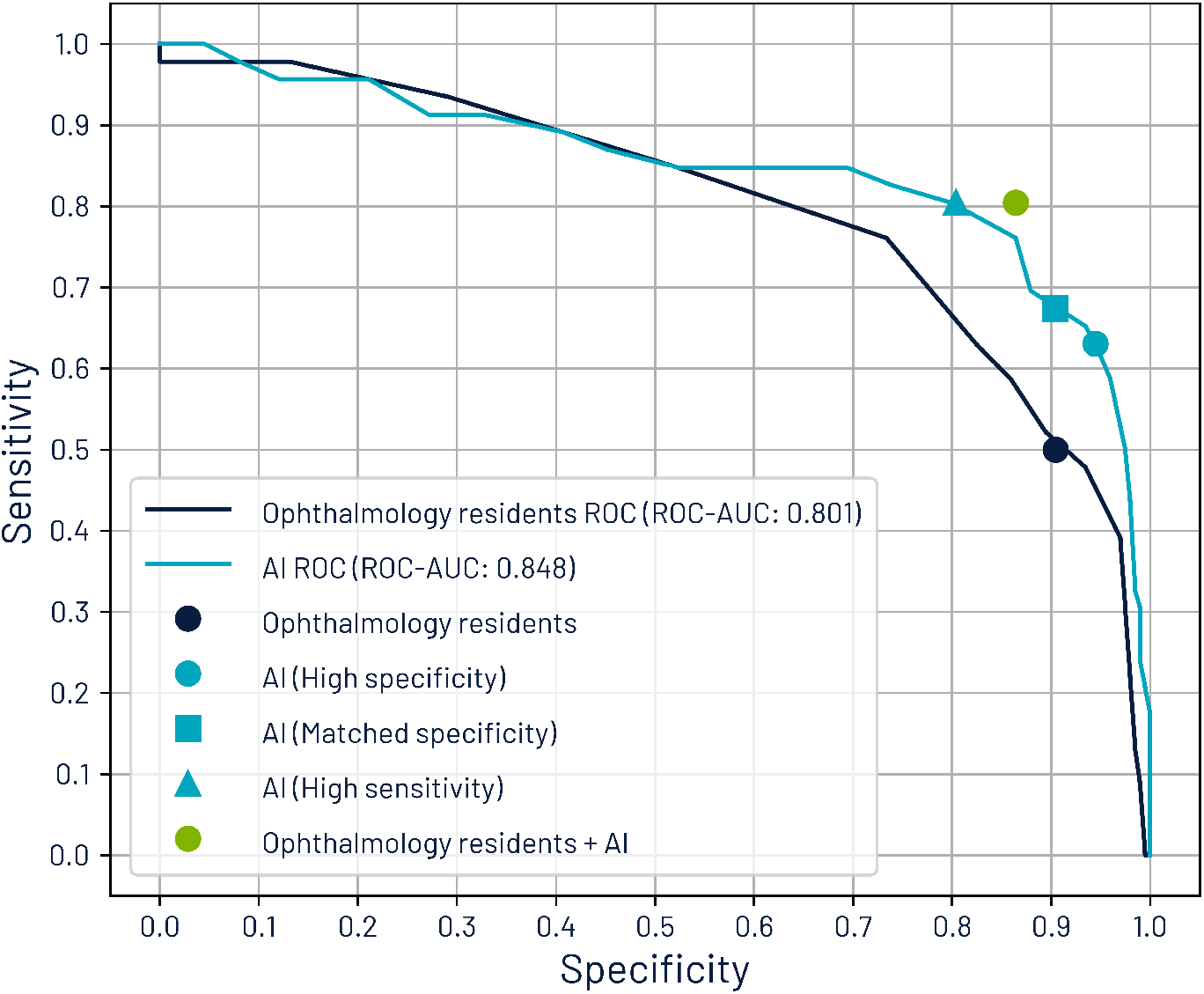
Receiver Operating Characteristic Curves for Glaucoma Suspect Classification Based on Maximum CDR by patient.

The area under the ROC (AUC-ROC) for AI is 0.848, while the AUC-ROC for residents of ophthalmology is 0.801.

We marked different operating points. One for sensitivity and specificity based on AI’s results (high specificity point and best F1-score), another for residents’ diagnoses, and one for the synergistic approach. Additionally, we mark the point where AI on its own matches the sensitivity of the combined approach attaining a sensitivity of 80.4% and specificity of 80.4%, and the point where AI’s specificity matches that of the resident’s, yielding a sensitivity of 67.4% and a specificity of 90.5%.

We further analysed CDR estimates for the 362 eyes evaluated by all 3 ophthalmologists, by the residents of ophthalmology and by the AI tool. The mean absolute error for CDR estimates was 0.056 (SD: 0.042) for AI and 0.105 (SD: 0.074) for residents. While the relative errors were 10.9% (SD: 7.9%) and 20.8% (SD: 14.4%) respectively. Performance differences were statistically significant with p < 0.001.

In Figure 2, we show the comparison between CDR ground truth values and CDR estimates for both AI and ophthalmology residents. For both, there is a significant relationship between estimation and ground truth (p < 0.001). However, the Pearson correlation coefficient for AI was higher (r = 0.728) than for ophthalmology residents Even though the AI tool provided a better CDR estimation, and the synergistic approach yielded better results than residents on their own, only 61.6% of patients were evaluated with AI due to image quality limitations.

**Figure 2:**
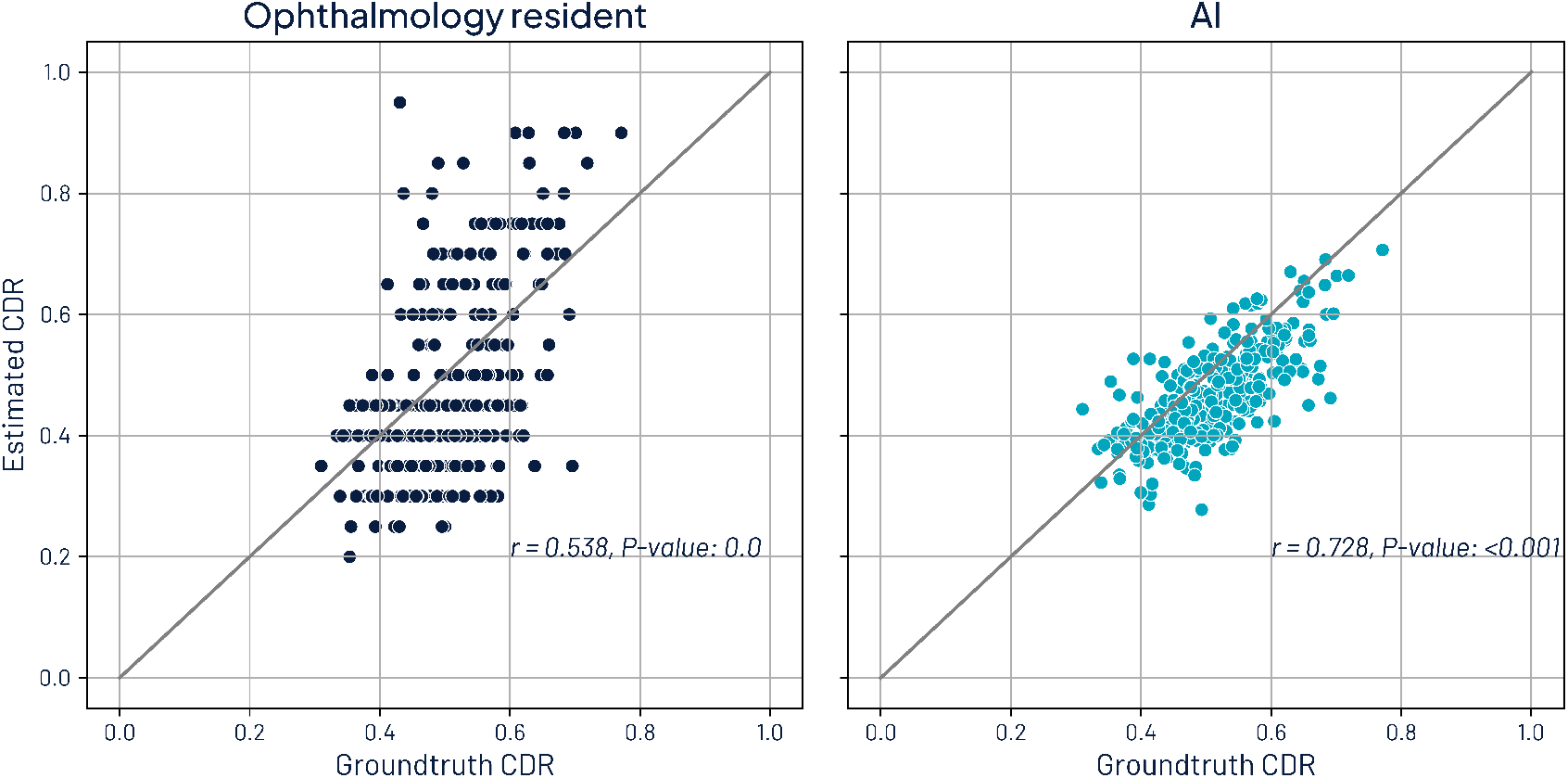
Comparison of CDR estimates with groundtruth CDR. The left image shows the comparison with the residents and the right one with AI.

To better understand the reasons related behind insufficient image quality, we compared the prevalence of residents’ diagnoses between the subset with ground truth CDR (78.6%) and the subset without (21.4%). Diagnoses where prevalence on the subset without ground truth exceeded by at least a 25% relative difference the subset with ground truth were PM (6.7% vs 0.5%), retinal detachment (10.0% vs 1.2%), vitreous haemorrhage (7.5% vs 1.5%), uveitis (0.8% vs 0.2%), non-functional eye or prosthesis (5.8% vs 1.5%), AMD (1.7% vs 0.5%), DR (5.8% vs 4.4%), and cataracts (9.2% vs 7.2%). From the latter, we may notice that opacities arising from retinal detachment, vitreous haemorrhage, and cataracts may be related to a decreased image quality. Moreover, high myopia (related to PM) also affects image quality resulting in blurry images.

### Retinal disease

The analysis was performed on 395 patients (90.8%) where at least one eye was evaluated by AI to determine presence of retinal disease. If the AI tool indicated presence of DR, ME, AMD, PM, or other retinal disease, it was considered positive for presence of retinal disease.

Table 2 presents performance metrics. We distinguished three categories for sensitivity analysis. The first category included the presence of all retinal disease, including those associated with mild stages of retinal disease (excluding tessellated fundus). The second category is associated with medium or high risk of vision loss and mild diseases are excluded. The third category corresponds to high risk of vision loss and encompasses findings such as vitreous haemorrhages, retinal detachment, and neovascularization. A complete list of what is included in each category is provided in Appendix D.

**Table 2:** Performance metrics for presence of retinal disease.

|  | Accuracy | Sensitivity | Sensitivity<br>medium risk<br>and over | Sensitivity<br>high risk | Specificity | PPV | F1 score |
| --- | --- | --- | --- | --- | --- | --- | --- |
| **Resident**<br>**diagnosis** | 75.2% (70.9%,<br>79.7%) | 51.9% (44.0%,<br>59.9%) | 63.0% (52.2%,<br>73.3%) | 80.5% (67.9%,<br>92.1%) | 90.4% (86.4%,<br>93.9%) | 77.9% (70.0%,<br>85.4%) | 62.3% (54.9%,<br>69.0%) |
| **AI** | **88.1%**<br>(84.8%,<br>91.1%) | 76.3% (69.9%,<br>82.6%) | 90.1% (83.5%,<br>96.1%) | **100.0%**<br>(100.0%,<br>100.0%) | **95.8%**<br>(93.1%,<br>98.3%) | **92.2%**<br>(87.3%,<br>96.5%) | **83.5%**<br>(78.7%,<br>87.6%) |
| **Resident**<br>**or AI** | 86.1% (82.8%,<br>89.4%) | **84.0%**<br>(78.4%,<br>89.9%) | **92.6%**<br>(86.5%,<br>97.7%) | **100.0%**<br>(100.0%,<br>100.0%) | 87.4% (82.9%,<br>91.2%) | 81.4% (75.3%,<br>86.7%) | 82.6% (78.1%,<br>86.9%) |

Sensitivity increases for both AI and ophthalmology residents as the risk of vision loss increases. Across all categories, AI’s sensitivity was statistically significantly better than that of the residents (p<0.001). For all retinal diseases AI achieved a sensitivity of 76.3%, outperforming residents at 51.9%. For retinal disease associated to medium or high-risk, AI’s sensitivity was 90.1% compared to residents’ 63.0%, and for high-risk, AI reached 100% sensitivity versus 80.5%. Significant differences were observed across all metrics. Performance metrics are shown on Figure 3.

**Figure 3:**
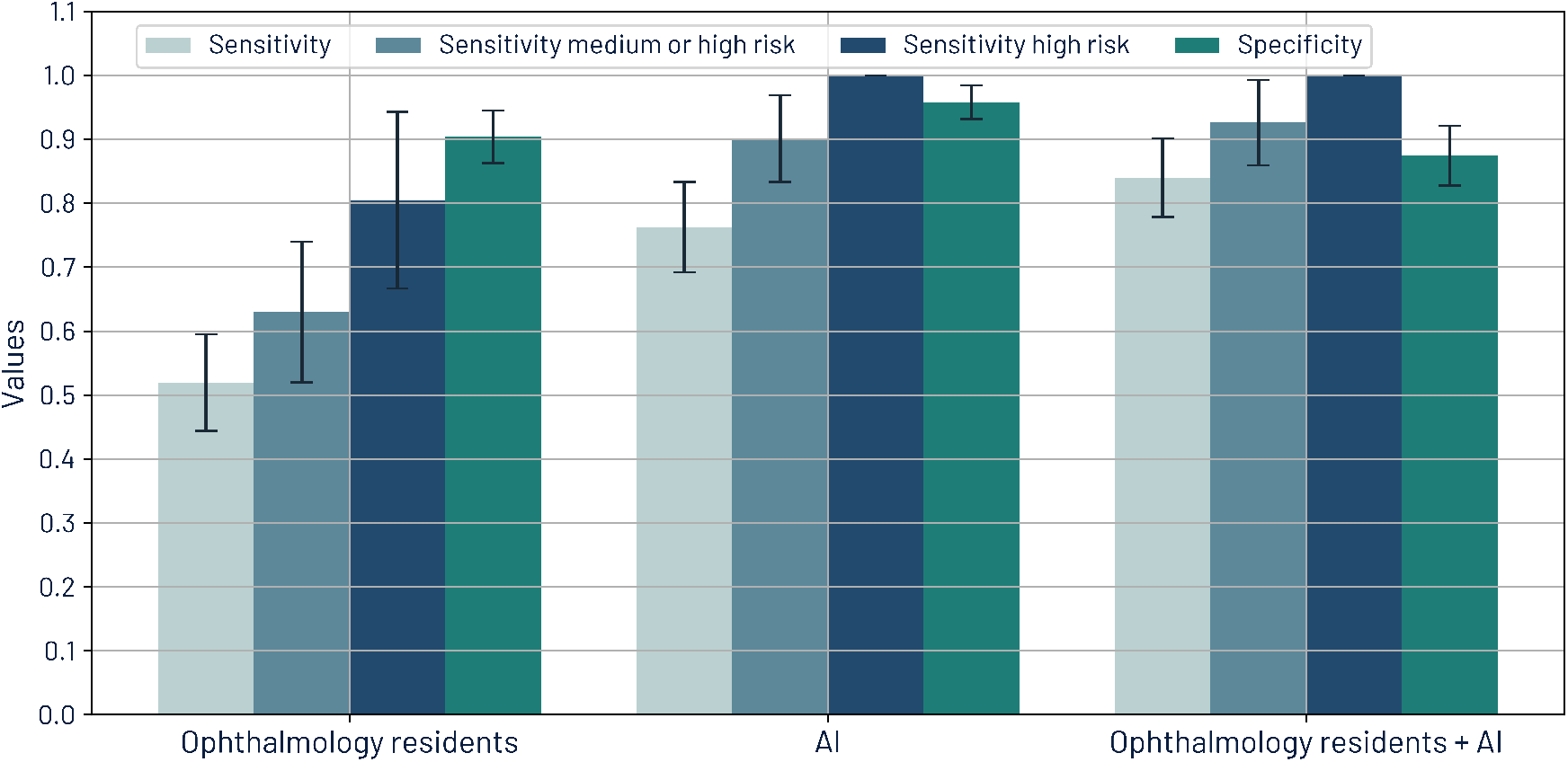
Sensitivities by risk of visual loss and specificity for retinal diseases for ophthalmology residents, AI, and the synergistic approach.

The synergistic approach led to higher sensitivities resulting in 84.0% for all retinal disease, 92.6% for medium or high risk, and 100% for high-risk. PPV for this approach is 81.4%, which is better than the baseline PPV of the residents’ diagnoses.

## DISCUSSION

In this study we evaluated the performance of an AI tool, the initial diagnosis of ophthalmology residents, and a combined approach for retinal disease analysis, CDR estimation and glaucoma suspect classification.

The AI tool demonstrated better performance compared to ophthalmology residents in CDR estimations, obtaining a lower average error (0.056 vs 0.105, p<0.001) and a higher Pearson correlation coefficient (0.728 vs 0.538). This difference was expected, since CDR estimation is challenging even for experts [17, 20]. Consequently, implementing this AI tool would potentially standardize CDR measurements. We did not assess how explainability features from the AI tool could help residents determine CDR measurements. This can be evaluated with further research to assess the benefits of having a visual aid to determine CDR.

Determining presence of glaucoma is highly related to OD assessments [29]. Furthermore, it has been shown that CDR is a significant indicator for presence of glaucoma [31]. Therefore, a better CDR estimation results in better glaucoma suspect classification. In this work we considered as glaucoma suspects, patients whose CDR *≥* 0.6 or for whom CDR difference between both eyes was larger than 0.2.

Under these criteria, sensitivity was higher for AI (63.0%), than for ophthalmology residents (50.0%). While both sensitivities are relatively low when considered independently and not statistically significantly different (p=0.116), when considering the synergistic approach sensitivity increased to 80.4% (p<0.001) with a specificity of 86.4%, suggesting the relevance of a combined approach between AI and residents in clinical practice.

For retinal disease, there is an important difference on sensitivity between AI and residents. For retinal disease with medium or high risk of visual loss sensitivity was 90.1% for AI compared to 63.0% for residents. Even when sensitivity increased for high-risk findings, residents only achieved 80.5% compared to a 100% by AI. Specificity is also higher for AI, resulting on an overall better evaluation from AI. Furthermore, AI was statistically significantly better across all metrics. Although not all patients require referral to further tests or to retina specialists, high sensitivity is important to make a better decision in the referral process and to provide better recommendations in terms of self-care and follow-up.

Implementing the AI tool demonstrates significant potential in enhancing sensitivity in retinal disease assessments and guiding more accurate measurements of CDR. However, it is crucial to emphasize that the final diagnosis must always be based on the ophthalmologist’s comprehensive evaluation.

Ophthalmologists perform a thorough assessment that encompasses examining the peripheral retina (which may not be visible in digital images), evaluating anterior segment structures, and identifying opacities that could compromise retinal image quality. These aspects of clinical examination are critical and cannot be fully replicated by current AI technologies. Furthermore, the decision-making process involving referral to a subspecialty, requesting additional studies or not referring at all, remains a critical aspect of the resident’s role.

Moreover, it must be considered that due to image quality, AI only estimated CDR for 61.6% of patients. For OD, poor image quality is a downside, which could be resolved by relaxing quality criteria, since all three ophthalmologists deemed 78.6% of patients had images with sufficient quality. To further increase evaluation, another option is using fundus cameras with higher resolution, however, this may also result in higher implementation costs. However, quality for OD evaluation may still be an issue for patients with opacities or high myopia due to physiological differences that difficult obtaining high quality images.

Nevertheless, AI can serve as a valuable tool to enhance sensitivity on retinal disease assessments, homogenize CDR estimates, and guide the selection of additional examinations, referral to subspecialty and follow-up. Moreover, a synergistic approach between AI and residents showed enhanced sensitivity, with little effect on specificity. Therefore, its use should be considered in future clinical practice.

Future research should focus on integrating AI tools into clinical workflows, evaluating their impact on patient outcomes, and exploring their potential in ophthalmology resident training programs.

## Data Availability

All data produced in the present study are available upon reasonable request to the authors.

## ACKNOWLEDGMENTS

The authors acknowledge Diana González and Nayelli Cruz for their contributions to project definition and coordination of the optometry team. Iris Pantoja, Brenda Camarillo, and Rodolfo Pineda are recognized for their work in conducting the AI screening. The authors also acknowledge Vanessa Tirado for annotating optic disk images.

