## Supplementary material for "Comparative Performance of an AI Tool and First-Year Residents for Retinal Disease and Glaucoma Assessments: A Study in a Mexican Tertiary Care Setting"

### A Variables collected during the screening process

List of variables collected for this research study.

| Code | Definition | Type | Values |
|---|---|---|---|
| expediente | Medical record ID. | text | - |
| patient_age | Age. | numeric | 0-120 |
| postalCode | Zip Code | text | - |
| gender | Sex assigned at birth. | categorical | 1: Male<br>2: Female |
| diabetes | Diagnosed diabetes. | boolean | 0: No<br>1: Yes |
| diabetesYears | Time with diagnosed diabetes. | numeric | 0-25 |
| hypertension | Diagnosed hypertension | boolean | 0: No<br>1: Yes |
| hypertensionYears | Time with diagnosed hypertension. | numeric | 0-25 |
| lastVisitOphthamologist | Date of last ophthalmic evaluation | categorical | 1: Within the last 12 months<br>2: More than a year ago<br>3: Never |
| sight_loss_in_family | Family history of vision loss | boolean | 0: No<br>1: Yes |
| visionAffectingQualityLife | Patient perceives that visual conditions interfere with daily activities | boolean | 0: No<br>1: Yes |
| SY-OPH-CAT-1 | Blurry or foggy vision even with glasses | boolean | 0: No<br>1: Yes |
| SY-OPH-CAT-2 | Changes in color perception | boolean | 0: No<br>1: Yes |
| SY-OPH-CAT-3 | Increased sensitivity to light or halos | boolean | 0: No<br>1: Yes |
| SY-OPH-CAT-4 | Difficulty to see at night | boolean | 0: No<br>1: Yes |
| SY-OPH-OT-2 | Perceived flashes or light streaks | boolean | 0: No<br>1: Yes |
| SY-OPH-OT-3 | Eye pain | boolean | 0: No<br>1: Yes |
| SY-OPH-OT-4 | Spots or patches that persist within vision all the time | boolean | 0: No<br>1: Yes |
| SY-OPT-PR-1 | Difficulty seeing up close or reading small print | boolean | 0: No<br>1: Yes |
| symptomDevelop | How did the changes in the patient's vision appeared. | categorical | 1: Suddenly<br>2: Progressively |

### B Models and architecture

#### B.1 Retinal disease assessment

For retinal disease analysis, retinIA incorporates a multi-output convolutional neural network to determine image quality, laterality, presence of diabetic retinopathy, macular edema, macular degeneration, pathological myopia, presence of retinal lesions and associated risks of vision loss.

Prior to implementing the model, images are resized to 512x512 and preprocessed using a method based on Graham [1].

The convolutional base corresponds to that of the Inception V3 architecture [2]. This is followed by a dense layer of 2,048 units, that further divides into two branches: one for image quality and laterality, and another for disease analysis and associated retinal lesion severity. Each output is evaluated independently for classification. Thus, it is an architecture with multiple classification outputs.

The complete architecture can be seen on Figure 1.

The model was trained on Mexican data with 104,216 fundus images, using 91,884 for training, 6,156 for validation and 6,176 for internal testing previous to this study.

These images were taken with different fundus non mydriatic cameras such as Horus 45° autofocus portable fundus non-mydriatic camera from Jedmed, Visuscout 100 from Zeiss, and DRS from Centervue. All images were annotated by a retina expert or an ophthalmologist to determine groundtruth values.

This AI tool also includes explainability features for the AI analysis performed on retinal images. These features include a colormap for retinal anomalies, obtained by combining GradCam [3] and SmoothGrad [4] methods. Figure 2 shows an example of image preprocessing for model input, and the explainability heatmap that arises from postprocessing.

#### B.2 CDR estimation

CDR estimation involves several steps. First, images are resized to 256x256, and a U-Net architecture [5] is used to segment the optic disk and identify the region of interest (ROI) where the optic disk is present. The optic disk ROI is extracted and resized to 256x256. Image quality is then verified using a convolutional neural network based on Inception V3 with 256x256 input. If quality meets the criteria, a second U-Net is used to extract the optic cup. The segmented image is re-scaled to the original ROI size, and the heights of the optic cup and optic disk are extracted to calculate CDR.

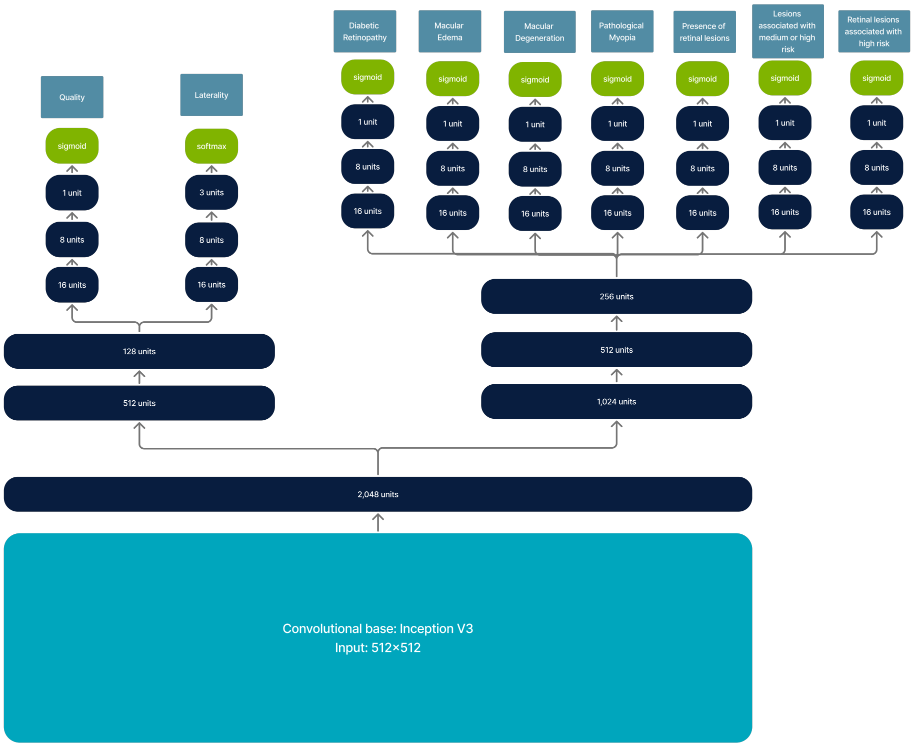


Figure 1: Architecture used for retinal image analysis

A close-up of the optic disk is provided by retinIA, with markings of the optic disk and optic cup heights and the CDR estimate.

Figure 3 shows graphically the process to estimate CDR, the final image corresponds to the explainability output provided by retinIA.

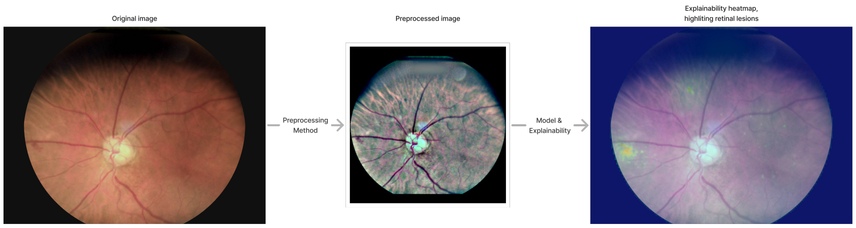


Figure 2: Indicates how the original image is preprocessed, and shows explainability heatmap generated by the AI tool

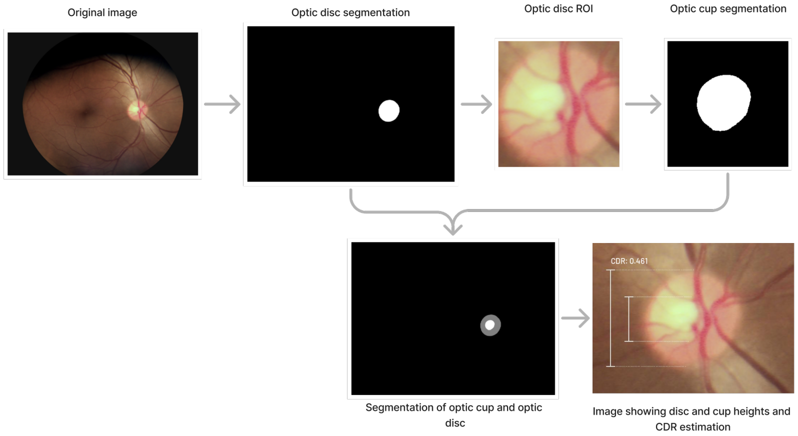


Figure 3: Process involved to determine CDR estimation.

The optic cup and optic disk segmentation models were trained on 18,446 images, 15,477 for training, 1,490 for validation, and 1,479 for internal testing. Corresponding masks were annotated by an ophthalmologist. As well as the model for retina assessment, images were taken with different fundus non mydriatic cameras such as Horus 45° autofocus portable fundus non-mydriatic camera from Jedmed, Visuscout 100 from Zeiss, and DRS from Centervue.

##### B.3 Media opacities

For media opacity assessment, the platform considers the following variables: image quality (resulting from the retina analysis), patient age, and cataract related symptoms, including blurry vision, changes in color perception, increased sensitivity to light and difficulty to see at night. Logical rules have been defined aided by ophthalmologists to determine possible media opacities. As an example, if a patient is 60 years or older, has blurry vision, and none of the images taken had enough quality for retina evaluation, the patient is classified as having possible media opacities.

### C Retinal findings annotated in fundus images

Retinal findings that were annotated by ophthalmologists are shown in Table 1.

| **Category** | **Findings** |
|---|---|
| Atrophies | S-AT-1: Diffuse chorioretinal atrophy<br>S-AT-2: Patchy chorioretinal atrophy<br>S-AT-3a: Geographic or macular atrophy involving the fovea<br>S-AT-3b: Geographic or macular atrophy not involving the fovea |
| Drusen | S-DR-1a: Less than 20 small drusen<br>S-DR-1b: 20 or more small drusen<br>S-DR-2a: Less than 20 medium drusen<br>S-DR-2b: 20 or more medium drusen<br>S-DR-3: Large drusen |
| Exudates | S-EX-1a: Hard exudates within one disk diameter of macula center.<br>S-EX-1b: Hard exudates further than one disk diameter from the macula center, but within the temporal arcades<br>S-EX-1c: Hard exudates outside of the temporal arcades<br>S- EX-2: Soft exudates |
| Hemorrhages and Microaneurysms | S-HM-1: Microaneurysms<br>S-HM-2: Vitreous hemorrhage<br>S-HM-3: Preretinal hemorrhage<br>S-HM-4: Superficial hemorrhage<br>S-HM-5a: Less than 20 intra-retinal hemorrhages<br>S-HM-5b: 20 or more intra-retinal hemorrhages<br>S-HM-6: Subretinal hemorrhage |
| Vascular anomalies | S-VA-1: Intraretinal microvascular abnormalities<br>S-VA-2: Venous beading<br>S-VA-3: Neovascularization<br>S-VA-4: Changes in the arteriolar light reflex<br>S-VA-5: Arteriovenous crossings<br>S-VA-6: Focal narrowing of retinal arterioles<br>S-VA-7: Generalised narrowing of retinal arterioles<br>S-VA-X: Other microvascular abnormalities |
| Other retinal findings | S-OT-1: Retinal pigmentary epithelium changes<br>S-OT-2: Choroidal neovascular membrane<br>S-OT-4: Lacquer cracks<br>S-OT-5: Fuch's spot<br>S-OT-6: Disciform scar<br>S-OT-7: Epiretinal membrane<br>S-OT-8: Macular hole<br>S-OT-9: Rhegmatogenous retinal detachment<br>S-OT-10: Tractional retinal detachment<br>S-OT-11: Central or branch artery occlusion<br>S-OT-12: Central or branch vein occlusion<br>S-OT-XR: Other pathological signs in retina |
| Non pathological findings | S-NP-1: Tessellated fundus<br>S-NP-2: Retinal fiber layer myelination<br>S-NP-3: Choroidal nevus<br>S-NP-4: Panretinal photocoagulation traces<br>S-NP-X: Other non-pathological findings |

Table 1: Findings annotated during the labeling process.

### D Logical rules to determine prediagnosis and risk of visual loss from findings in fundus images

To define DR stages we used the International Clinical Diabetic Retinopathy and Diabetic Macular Edema Disease Severity Scales (ICDR) [6].

According to the Guidelines on Diabetic Eye Care, DME is defined as retina thickening, however this cannot be measured on a 2D fundus images. [7] Thus, we use hard exudates as a proxy of DME, since they are a sign of current or previous DME. [7] Moreover, Litvin *et al* found that detecting hard exudates within one disc diameter (DD) from the fovea had a 93.8% sensitivity for clinically significant DME.[8] Therefore, we consider hard exudates within one DD as a proxy for center-involving diabetic macular edema, and other hard exudates within the macula area as a proxy for non-center-involving diabetic macular edema.

For AMD there are several definitions and severity scales, such as [9, 10, 11, 12, 13, 14, 15, 16]. Most of them consider drusen, retinal pigmentary epithelium (RPE) changes, geographic atrophy; and choroidal neovascularization as well as other signs of neovascular maculopathy such as RPE detachment. We considered a simplified version of the Preferred Practice Pattern (PPP) scale used by Flaxel *et al* [16]. We considered the mild stage to have more than 20 small drusen, from 1 to 19 medium drusen, or RPE changes. The moderate stage included more than 20 medium drusen, at least one large druse, or non-central geographic atrophy. For the advanced stage we considered either geographic atrophy involving the foveal center or a choroidal neovascular membrane additional to drusen or RPE changes.

To classify pathological myopia we used the categorization by Ohno-Matsui *et al* considering the 4 stages and an additional stage if any of the "plus" lesions was present.[17]

To determine which retinal findings corresponded to the different levels of risk of visual loss, we consulted two retina specialists. Findings on the medium or high risk category, correspond to those for which both retina experts agreed a consultation with a retina expert was required. Findings on the high risk of visual loss, were those that could require treatment. The latter, considering hard exudates on the macular area as a proxy for macular edema, and excluding vitamin prescription for AMD within the treatments.

The corresponding rules that determine possible prediagnosis and risk of visual loss are presented on Table 2.

| Disease | Severity | Rules |
|---|---|---|
| Diabetic Retinopathy | Mild | S-HM-1 |
| | Moderate | (S-HM-1 **AND** (S-EX-1a **OR** S-EX-1b **OR** S-EX-1c **OR** S-EX-2)) **OR** S-HM-5a |
| | Severe | ((S-HM-1 **OR** S-HM-5a) **AND** (S-VA-1 **OR** S-VA-2)) **OR** S-HM-5b |
| | Proliferative | S-VA-3 **OR** S-HM-2 **OR** S-HM-3 |
| Macular Edema | Non-central | S-EX-1b |
| | Central | S-EX-1a |
| Age Macular Degeneration | Mild | S-DR-1b **OR** S-DR-2a **OR** S-OT-1 |
| | Intermediate | S-DR-2b **OR** S-DR-3 **OR** (S-AT-3b **AND** (S-DR-1 **OR** S-DR-2a **OR** S-OT-1)) |
| | Advanced | (S-AT-3a **OR** S-OT-2) **AND** (S-DR-1 **OR** S-DR-2a **OR** S-DR-2b **OR** S-DR-3 **OR** S-OT-1) |
| Pathological Myopia | 1 | S-NP-1 |
| | 2 | S-NP-1 **AND** S-AT-1 |
| | 3 | S-NP-1 **AND** S-AT-2 |
| | 4 | S-NP-1 **AND** (S-AT-3a **OR** S-AT-3b) |
| | PLUS | S-NP-1 **AND** (S-OT-2 **OR** S-OT-4 **OR** S-OT-5) |
| Retinal findings | - | S-VA-3<br>**OR** S-AT-1 **OR** S-AT-2 **OR** S-AT-3a **OR** S-AT-3b<br>**OR** S-DR-1b **OR** S-DR-2a **OR** S-DR-2b **OR** S-DR3<br>**OR** S-EX-1a **OR** S-EX-1b **OR** S-EX-1c **OR** S-EX-2<br>**OR** S-HM-1 **OR** S-HM-2 **OR** S-HM-3 **OR** S-HM-4 **OR** S-HM-5a **OR** S-HM-5b **OR** S-HM-6<br>**OR** S-OT-1 **OR** S-OT-2 **OR** S-OT-4 **OR** S-OT-5 **OR** S-OT-6 **OR** S-OT-7 **OR** S-OT-8 **OR** S-OT-9 **OR**<br>S-OT-10 **OR** S-OT-11 **OR** S-OT-12 **OR** S-NP-4 **OR** S-OT-XR |
| Medium or high risk of visual loss | - | S-VA-3<br>**OR** S-AT-2 **OR** S-AT-3a **OR** S-AT-3b<br>**OR** S-DR-2b **OR** S-DR3<br>**OR** S-EX-1a **OR** S-EX-1b<br>**OR** S-HM-2 **OR** S-HM-3 **OR** S-HM-5a **OR** S-HM-5b **OR** S-HM-6<br>**OR** S-OT-1 **OR** S-OT-2 **OR** S-OT-6 **OR** S-OT-8 **OR** S-OT-9 **OR**<br>S-OT-10 **OR** S-OT-12 **OR** S-NP-4 |
| High risk of visual loss | - | S-VA-3<br>**OR** S-EX-1a **OR** S-EX-1b<br>**OR** S-HM-2 **OR** S-HM-3 **OR** S-HM-6<br>**OR** S-OT-2 **OR** S-OT-8 **OR** S-OT-9 **OR**<br>S-OT-10 **OR** S-OT-12 |

Table 2: Severity grades for DR, ME, AMD, and PM and the lesions in retina that conform them. For lesions in retina we exclude vascular anomalies other than neovascularization (S-VA-3). We also excluded non-pathological findings, except for panretinal photocoagulation traces, which may require continuous assessment by a retina specialist.

### E Cataract and media opacity comparison

During the data collection process, information on cataract diagnosis and referral to cataract surgery was obtained. Also, retinIA's possible outcomes include presence of media opacity. Although there was no ground truth value for cataract, and currently retinIA does not have the capability to distinguish cataracts from other media opacities, we compared the residents' diagnoses with the results from the AI tool.

Percentage of agreement between the ophthalmology resident and the AI tool was 86.0%. Table 3 shows the confusion matrix that compares media opacity detection from retinIA and cataract diagnosis from ophthalmology residents.

| | **No media opacity (AI)** | **Media opacity (AI)** |
|---|---|---|
| **No cataract (resident)** | 360 | 35 |
| **Cataract (resident)** | 26 | 14 |

Table 3: Confusion matrix for comparing media opacity detection from retinIA and cataract diagnosis from ophthalmology residents.

Even though the percentage of agreement is high, the confusion matrix shows that there is little agreement on detection itself, which accounts for minimal agreement in terms of Cohen's Kappa ($\kappa = 0.237$) [18]. This low agreement may be due to the presence of opacities or cataract-related symptoms that correspond to different diseases. Also, presence of some cataracts still allow for fundus imaging with sufficient quality for analysis of the retina, and therefore may not be considered by the AI tool as opacities.
